# Development and Validation of Automated Software for the Detection of Large Vessel Occlusion on Noncontrast CT

**DOI:** 10.1101/2024.10.14.24315498

**Authors:** Hirofumi Tsuji, Akira Ishii, Yu Abekura, Hidehisa Nishi, Takuya Fuchigami, Atsushi Tachibana, Hirotaka Ito, Yoshiki Arakawa

**Affiliations:** Department of Neurosurgery, Kyoto University Graduate School of Medicine, Japan; Department of Neurosurgery, Shizuoka General Hospital, Japan; Department of Neurosurgery, Juntendo University Graduate School of Medicine, Japan; Department of Neurosurgery, Kokura Memorial Hospital, Japan; Medical Systems Research and Development Center, FUJIFILM Corporation, Japan

**Keywords:** acute ischemic stroke, automated detection, machine learning

## Abstract

**Background:** Reliable detection of large-vessel occlusion (LVO) via medical-image analysis has significant advantages in cases of acute ischemic stroke (AIS). In recent years, convolutional neural network (CNN)-based technologies for automated LVO detection have been developed. However, the pros and cons of CNN-based assistance in clinical practice remain poorly understood. The purpose of this study was to develop and validate a deep learning-based model to detect the hyperdense-artery sign (HAS) as a proxy for LVO and to investigate its impact on neurosurgeons’ diagnostic accuracy.

**Methods:** We conducted a multicenter, retrospective study of patients with LVO due to anterior-circulation AIS who underwent computed tomography angiography or magnetic resonance angiography on admission, as well as patients without LVO (patients with no indicative angiography features and patients with cerebral infarction without LVO), who were admitted from 2006 to 2022. A CNN algorithm for LVO detection was developed using data from four institutions (n=690), and model performance was validated using data from five institutions (n=129). For further investigation, five board-certified and five non-board-certified neurosurgeons performed two separate observer-performance studies with a 4-week interval, with and without the CNN for each image.

**Results:** The HAS was detected in the correct location with a sensitivity and specificity of 0.79 and 0.87 by the CNN, 0.61 and 0.60 by board-certified neurosurgeons, and 0.61 and 0.66 by non-board-certified neurosurgeons, respectively. With the CNN, the mean area under the curve and figure of merit significantly increased for all readers (from 0.72 to 0.81, p<0.001, and from 0.71 to 0.77, p=0.005, respectively).

**Conclusions:** Our deep learning-based automated LVO-detection model for non-contrast-enhanced computed tomographic images significantly improved neurosurgeons’ diagnostic performance. Further studies are needed to clarify the usefulness of the CNN in clinical practice.

## Introduction

Large-vessel occlusion (LVO) is the obstruction of large, proximal cerebral arteries and accounts for approximately 30% of cases of acute ischemic stroke (AIS).^1^ Advances in mechanical thrombectomy have played an important role in the improvement of LVO treatment, which has increased the importance of rapid and reliable LVO detection and diagnosis.

The gold standard to detect LVO is the performance of computed tomography (CT) angiography (CTA) after bleeding is ruled out with non-contrast-enhanced computed tomography (NCCT).^2^ However, if LVO can be detected with high accuracy using NCCT alone, many advantages can be expected. For instance, in primary stroke centers, which have no neuroradiologists or neuro-interventionalists on staff, such detection is expected to reduce the number of missed cases^3^ and shorten the time before patients are transported to comprehensive stroke centers (CSCs). In terms of prehospital triage, one group reported on the direct guidance of patients to an angiography suite via a mobile stroke unit equipped with a CT scanner,^4^ indicating that NCCT may provide useful information. Moreover, at CSCs, reliable detection of LVO upon NCCT may allow circumvention of CTA studies and direct transfer of the patient to the angiographic suite, thus reducing wasted time and contrast media.

In recent years, convolutional neural network (CNN) technologies have been developed for medical image analysis, including automated LVO-detection models based on CTA or NCCT,^1,5,6^ but the impact of CNN assistance in clinical practice is not yet fully understood. Of particular importance are the potential advantages and disadvantages of these artificially intelligent technologies for clinicians with different levels of stroke-care experience. In this study, we developed a unique CNN algorithm for automated LVO detection based on NCCT and investigated its impact on neurosurgeons’ image-reading performance.

## Methods

### Data availability

The data that support the findings of this study are available, upon reasonable request, from the corresponding author.

### Patient population

In this multicenter, retrospective study, we used data of patients with anterior-circulation AIS, both for an LVO-positive group and a control group. For the LVO-positive group, the inclusion criteria were as follows: AIS in the anterior circulation (intracranial portion of the internal carotid artery, M1 and M2 portions of the middle cerebral artery), CT and magnetic resonance imaging (MRI) performed before treatment, LVO upon angiography and attempted intravenous thrombolysis or mechanical thrombectomy, and hospital arrival within 24 h since last-known-well time. The exclusion criteria were a stroke in the posterior circulation and unavailability of pretreatment CT or MRI scans. We also excluded scans with artifacts or excessive noise and those indicative of a tumor. Finally, NCCT scans were excluded if two neuro-interventionalists agreed that the hyperdense-artery sign (HAS) could not be confirmed.

For the control (LVO-negative) group, patients with suspected AIS who underwent CT or MRI and were proven to have no LVO were included. This group included the non-LVO ischemia (lacunar stroke, distal M3/M4 stroke, and so on), intracranial hemorrhage, and non-stroke groups.

Seven stroke centers participated in the study. For the derivation cohort, we collected data from Kobe City Medical Center General Hospital from 2011 to 2021, Kokura Memorial Hospital from 2013 to 2018, Kyoto University Hospital from 2008 to 2020, and Saiseikai Kumamoto Hospital from 2006 to 2017. For the validation cohort, we collected data from Kobe City Medical Center General Hospital from 2013 to 2015, Kyoto Medical Center from 2018 to 2020, Kyoto University Hospital from 2012 to 2022, Rakuwakai Otowakinen Hospital in 2021, and Koseikai Takeda Hospital from 2013 to 2019.

The study was conducted in accordance with the guidelines of the Declaration of Helsinki and was approved by the Kyoto University Graduate School and Faculty of Medicine, Ethics Committee: R2509-3. Patient informed consent for participation in this study was obtained in an opt-out fashion.

### Image acquisition

The pretreatment CT images were obtained with eight different scanners (Philips Ingenuity Core, Philips iCT 256, Philips Brilliance 64, Siemens SOMATOM Edge Plus, Canon Aquilion, Canon Aquilion PRIME, Canon Aquilion ONE, and GE Medical Systems Optima CT660), and the pretreatment MRI scans were obtained with five different scanners (Siemens Avanto, Siemens Skyra, Siemens Symphony, Canon Vantage Titan, and GE Medical Systems Signa HDXt).

### Labeling

Each HAS on each NCCT image was manually segmented by two board-certified neuro-interventionalists (H.T. and Y.Ab.), referring to CTA or magnetic resonance angiography (MRA) images taken almost simultaneously with the NCCT image. When the labeling by the neuro-interventionalists differed, a decision was made by consensus. These neuro-interventionalists were blinded to the patients’ clinical information.

### HAS segmentation algorithm pipeline

Our pipeline was composed primarily of three steps: midline correction, ischemic-core segmentation, and HAS segmentation (Figure 1). First, each Alberta Stroke Program Early CT Score (ASPECTS) region on the CT image is segmented using a two-dimensional U-Net-based algorithm (U-Net is a CNN architecture for semantic segmentation, widely used in the medical field). Thereafter, the CT image is rotated and translated with reference to the brain’s midline, which is derived by calculating the perpendicular bisector of the line connecting the left and right centers of gravity of the ASPECTS regions. Second, the ischemic core is segmented using the three-dimensional, fully CNN-based brain-hemisphere-comparison algorithm.^7–9^ Finally, the HAS is segmented by inputting the midline-corrected image and the ischemic core region to a similar network as that used in step 2, noting that the location of the ischemic core is related to that of the HAS. Although the three-dimensional, fully CNN-based brain-hemisphere-comparison model was designed to calculate the ASPECTS, we also used it to perform ischemic-core and HAS segmentation because the key concept (deep learning-based left-right comparison) was considered useful for the delineation of early ischemic changes.

**Figure 1.**
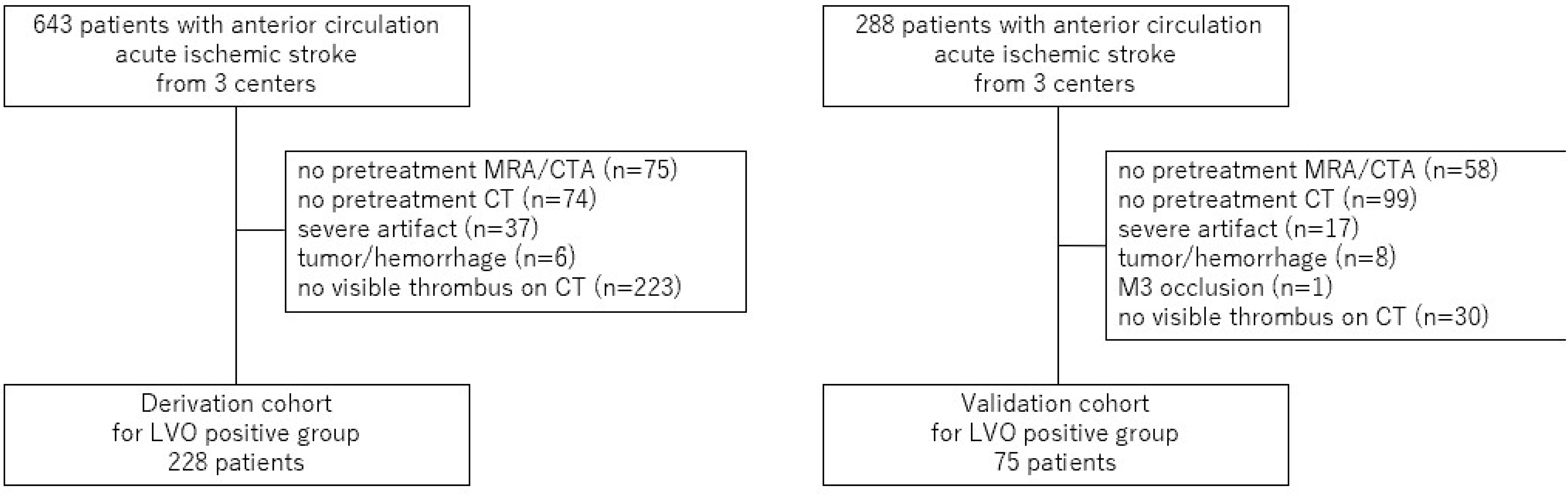
Algorithm pipeline for HAS segmentation. HAS, hyperdense-artery sign

### Model training and internal evaluation

For training of the HAS-segmentation network, the voxel spacing in the x and y directions of all CT images was rescaled to 0.75 mm, and that in z direction ranged from 3.00 to 8.00 mm. Cross-entropy loss was used as the loss function. In total, 60 CT images (approximately 10% of all the CT images of validation data) were randomly selected for internal validation, and the remaining data were used for training. The final model architecture and hyperparameters were chosen based on the internal validation results.

### Observer-performance study by neurosurgeons

To verify the effect of the CNN model on image-reading accuracy, 10 neurosurgeons (five board-certified and five non-board-certified neurosurgeons) were included in the observer-performance study. The board-certified neurosurgeons had 9–21 years of experience (mean: 15.4 years), and the non-board-certified neurosurgeons had 1–3 years of experience (mean: 1.8 years). The sequential testing method was used in the observer-performance study.^10^ All observers were blinded to the results of the consensus labeling by the two independent neuro-interventionalists. Validation data were randomly assigned for reading with and without the CNN for each observer, and observers were asked to mark only the one pixel most obviously reflecting the HAS per thrombus by referring to the NCCT only and rate their confidence level (0-100) for a potential lesion. In the case of reading with CNN assistance, the observer was required to mark the thrombus by themself while referring to the thrombus sites automatically segmented by the CNN. If more than two thrombi were observed, multiple thrombus sites were marked. If no thrombus was detected, the observer marked nothing, and their confidence level was rated as zero. After a 4-week interval, the reading session was repeated with the same validation data and assigned for reading with and without the CNN in a configuration opposite to that in the previous session. The observers were blinded to the number of patients with LVO and the performance level of the CNN. For each case, the reading time was recoded to investigate the difference in the performance time with or without CNN. Before conducting the observer-performance test, each observer underwent a training session with seven training cases to become familiar with the CNN output and the test procedure. The training cases were not used in the observer-performance test.

### Statistical analysis

Continuous variables were expressed as means ± standard deviations or medians with interquartile ranges, depending on the distribution of the variable. Categorical variables were expressed as frequencies and percentages. Two-sided p values of <0.05 were considered to indicate statistical significance. The performance for the detection of HASs was evaluated by using areas under the curve (AUC) obtained from receiver operating characteristic (ROC) analysis and figures of merit (FOMs) obtained from jackknife free-response ROC analysis.^11^ The statistical significance of differences between the mean AUCs or FOMs of any of the readers was estimated using paired Student’s t tests. All statistical analyses were performed with Python software (Python Software Foundation, Beaverton, OR).

## Results

### Baseline characteristics

Figure 2 is the flowchart of the selection of the LVO-positive group in this study. Of 643 and 288 eligible patients, 228 and 75 were included in the derivation and validation cohorts, respectively. Regarding the control group, as summarized in Table 1, the derivation cohort included 462 patients, and the validation cohort included 54 patients. As a result, the total number of patients was 690 in the derivation cohorts and 129 in the validation cohorts. The 60 cases used for internal validation were obtained from the derivation cohorts and were independent of the validation cohorts.

**Figure 2.**
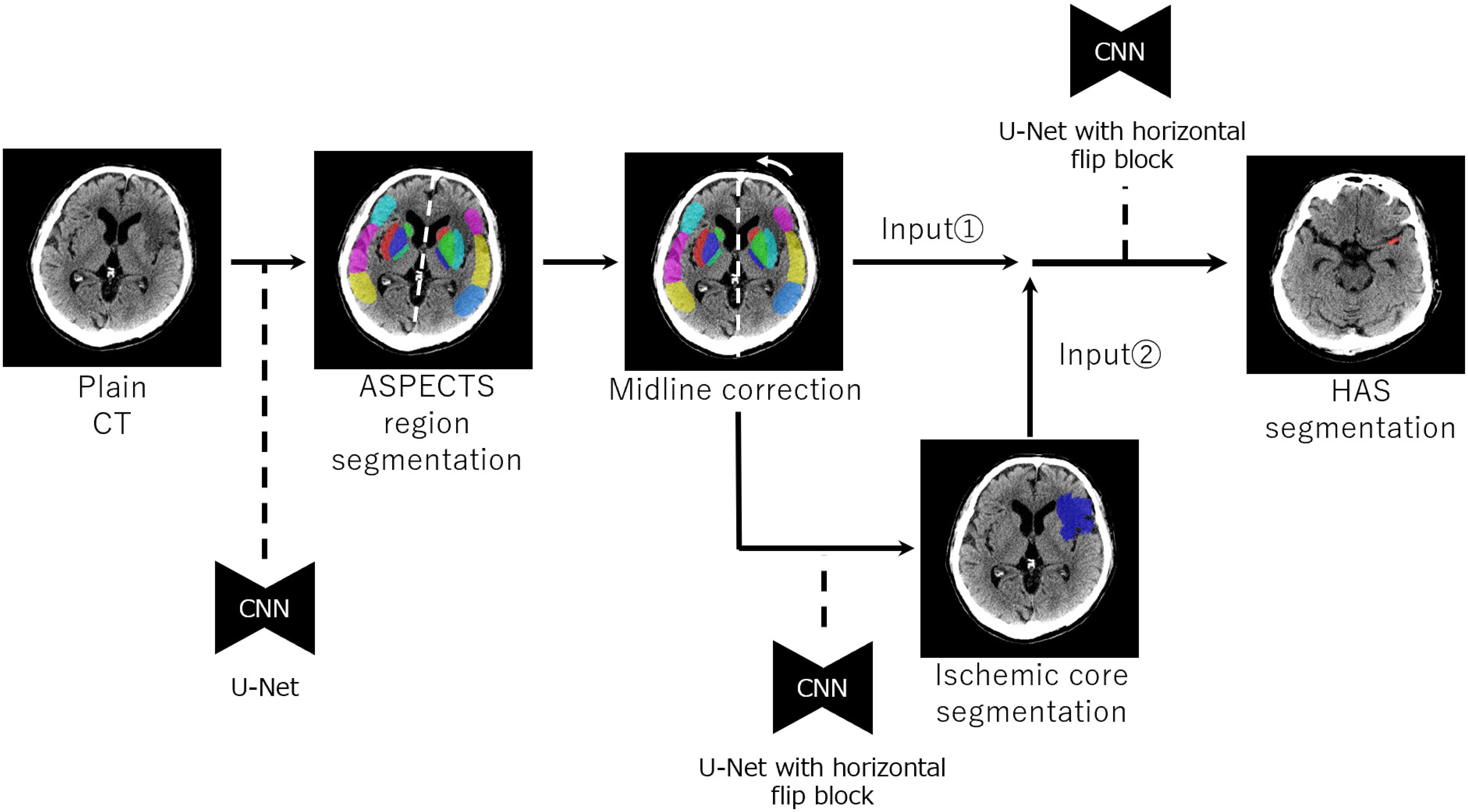
Flowchart of selection of the LVO-positive group included in this study. For all patients included in the LVO-positive group, intravenous thrombolysis or mechanical thrombectomy was attempted LVO, large-vessel occlusion

**Table 1.** Patient characteristics of each cohort.

|  | Derivation cohort | Validation cohort | P-value |
| --- | --- | --- | --- |
| Total number (n) | 690 | 129 |  |
| LVO positive, n (%) | 228 (33.0) | 75 (58.1) | <0.001 |
| Mean±SD age, years | 72±14.1 | 74±14.2 | 0.68 |
| Women, n (%) | 297 (43.0) | 60 (46.5) | 0.29 |
| LVO site |  |  |  |
| ICA, n (%) | 68 (29.8) | 21 (28.0) |  |
| M1, n (%) | 95 (41.7) | 42 (56.0) |  |
| M2, n (%) | 65 (28.5) | 12 (16.0) |  |
| Control group, n (%) | 462 (67.0) | 54 (41.9) |  |
| Non-LVO cerebral infarction, n (%) | 48 (10.4) | 34 (63.0) |  |
| Non-cerebral infarction, n (%) | 146 (31.6) | 20 (37.0) |  |
| Intracranial hemorrhage, n (%) | 268 (58.0) | 0 (0) |  |

ICA, internal carotid artery; LVO, large-vessel occlusion; SD, standard deviation

The derivation and validation cohorts did not significantly differ in terms of baseline characteristics except for the percentage of patients with LVO (Table 1). The derivation cohort comprised a larger proportion of patients without LVO. This was mostly attributed to the inclusion of patients with intracranial hemorrhage to improve the generalizability of the model. Regarding the validation cohort, we performed an observer study in terms of detection of LVO in a group of patients with cerebral infarction after exclusion of those with hemorrhage.

### Performance of the model and the observers

In 129 images in the validation cohort, the site of occlusion was the distal internal carotid artery in 21, the M1 segment in 42, and the M2 segment in 12 cases (Table 1). The HAS was identified in the correct location with a sensitivity and specificity of 0.79 and 0.87, respectively, by our model. In the observer study with the validation cohort, without CNN, the sensitivity and specificity were 0.61 and 0.60, respectively, by board-certified neurosurgeons, and 0.61 and 0.66, respectively, by non-board-certified neurosurgeons. With CNN usage, non-board-certified neurosurgeons exhibited a significant improvement in the AUC (from 0.73 to 0.80; p=0.023) but not in the FOM (from 0.71 to 0.75; p=0.10), whereas board-certified neurosurgeons exhibited significant improvements in both the AUC (from 0.71 to 0.83; p=0.005) and FOM (from 0.71 to 0.79; p<0.001) (Figure 3).

**Figure 3.**
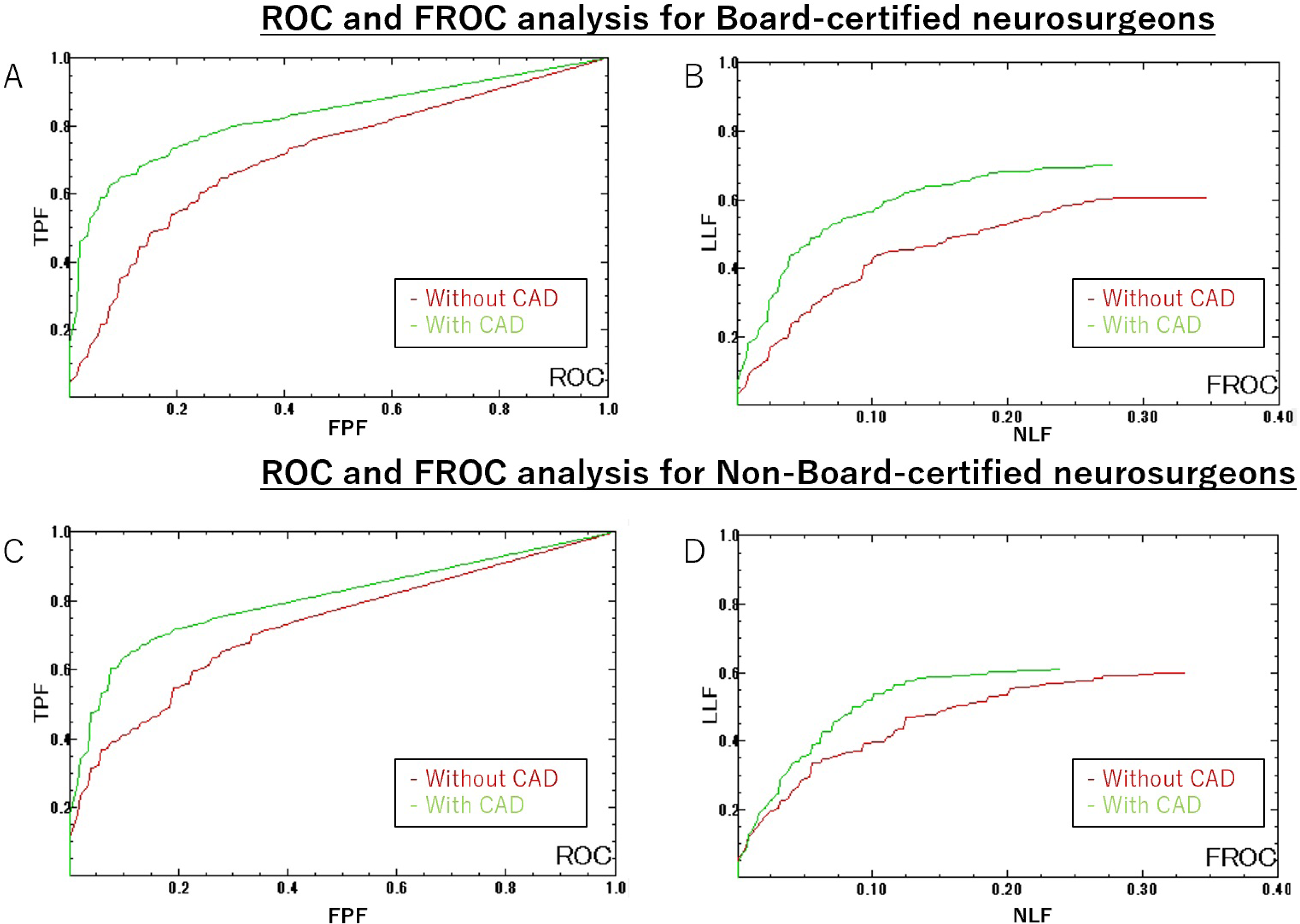
ROC and FROC analysis for neurosurgeons with/without CNN. (A) The AUC for board-certified neurosurgeons significantly improved (from 0.71 to 0.83). (B) The FOM for board-certified neurosurgeons also significantly improved (from 0.71 to 0.79). (C) The AUC for non-board-certified neurosurgeons significantly improved (from 0.73 to 0.80). (D) The FOM seemingly improved (from 0.71 to 0.75), but this was not significant. AUC, area under the curve; CNN, convolutional neural network; FOM, figure of merit; FROC, free-response receiver operating characteristic; ROC, receiver operating characteristic

For board-certified neurosurgeons, the reading time did not differ with the addition of CNN assistance (from 21.2 s to 20.6 s; p=0.48). However, for non-board-certified neurosurgeons, the reading time significantly increased with CNN assistance (from 19.4 s to 21.5 s; p=0.018).

The impact of CNN assistance on the reading performance of neurosurgeons in representative cases is depicted in Figure 4. Regarding the CNN’s positive effect, in the LVO-positive group, the use of the CNN changed the decision from a false negative to a true positive in 25 of 75 cases (33.3%) among board-certified neurosurgeons and in 17 cases (22.7%) among non-board-certified neurosurgeons. Moreover, in the LVO-negative group, the use of the CNN changed the decision from a false positive to a true negative in 32 of 54 cases (59.3%) among board-certified neurosurgeons and in 29 cases (53.7%) among non-board-certified neurosurgeons. However, the CNN also had a negative impact on neurosurgeons’ reading accuracy in some instances. In the LVO-positive group, the use of the CNN changed the decision from a true positive to a false negative in 6 of 75 cases (8.0%) among both board-certified and non-board-certified neurosurgeons. Moreover, in the LVO-negative group, the use of the CNN changed the decision from a true negative to a false positive in 6 of 54 cases (11.1%) in both groups of neurosurgeons.

**Figure 4.**
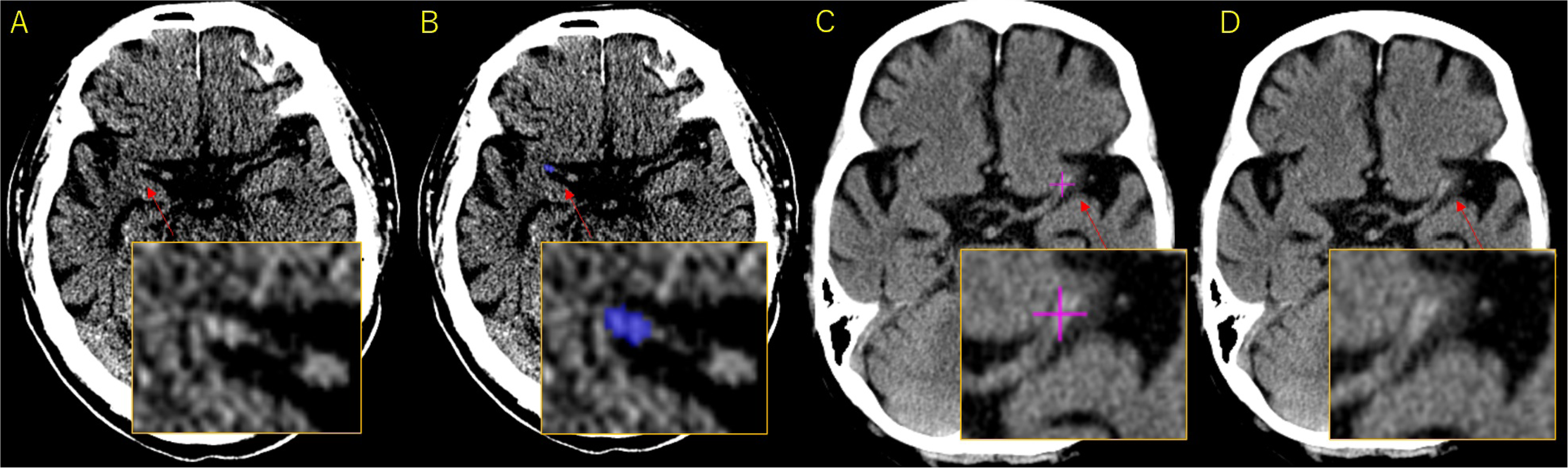
The effect of CNN assistance on reading performance for neurosurgeons in representative cases. (A) Original NCCT image with thrombus signals missed by the observer, and (B) correct segmentation achieved with the CNN (blue mark). With the assistance of the CNN, the observer’s diagnosis was corrected from a false negative to a true positive. (C) Original NCCT image incorrectly marked by the observer with a thrombus signal (pink crossbar), and (D) original NCCT not marked with the CNN. With CNN assistance, the observer’s diagnosis was corrected from a false positive to a true negative CNN, convolutional neural network; NCCT, non-contrast computed tomography

## Discussion

In this study, we developed a machine learning-based LVO-detection model for NCCT images. In the observer study, this model significantly improved the AUC performance of neurosurgeons. The proposed model is fully automated, requires no preprocessing, and processes an image in about 2.95 s.

In developing the model, we prioritized the detection of LVO with a high degree of confidence on NCCT images, to skip CTA or MRI in the clinical workflow. We minimized the false-positive rate by prioritizing specificity over sensitivity in the model. This approach was demonstrated not only in the performance of the CNN alone but also in the improvement of the clinicians’ accuracy in the LVO-negative group in the observer study.

Several reports on the performance of commercially available or original, automated LVO-detection models using only NCCT have been published.^12–15^ In a recent article, Weyland et al. reported the sensitivity and specificity achieved by using an original model in a non-consecutive cohort of patients with AIS.^12^ In their study, among 154 patients, 84 (54.5%) had CTA-confirmed LVO. The HAS was detected on the correct side with a sensitivity and specificity of 0.77 (95% CI, 0.66–0.85) and 0.87 (0.77–0.94) by the software, respectively. Olive-Gadea et al.^14^ reported the validation results of a commercially available automated software (MethinksLVO; Methinks Software S.L., Barcelona, Spain) in the identification of LVO on NCCT images. Among 1453 patients, 823 (56.6%) had LVO upon CTA. The HAS was detected in the correct location with a sensitivity of 0.83 and specificity of 0.71, and the AUC for LVO identification was 0.87. When the National Institutes of Health Stroke Scale (NIHSS) score upon admission was provided to the model, the sensitivity, specificity, and AUC improved to 0.83, 0.85, and 0.91, respectively. Validation data in these reports were mostly based on non-consecutive cases, impeding direct comparisons. However, as our model, which emphasizes specificity over sensitivity, had comparable accuracy to previously reported models, we believe that it is reliable. Achieving higher accuracy in imaging studies without contrast media would require the incorporation of additional clinical information, such as the NIHSS score,^14^ but this would be a trade-off against the complexity of the time and effort required in the clinical workflow, whether in the prehospital triage or the mobile stroke unit.

The following two points are possible advantages of automatic HAS segmentation on NCCT images: 1) it utilizes the characteristics of the HAS, and 2) it reduces the total volume of contrast agent used. Regarding the first advantage, the HAS is thought to represent the intravascular thrombus,^16^ and the red blood cell (RBC) composition of the thrombus is positively related to the CT value and the likelihood of identification of an HAS. Although no relationship between the RBC/fibrin composition of the thrombus and stroke etiology has yet been discovered,^17^ several reports have confirmed that RBC-rich thrombi are associated with high response rates to tissue plasminogen activator and thrombectomy.^18^ Regarding the second advantage, in the emergency department, physicians are hesitant to use contrast agents in patients with a background of kidney dysfunction or allergies and in patients for whom such clinical background information is difficult to obtain owing to aphasia or impaired consciousness. In AIS, baseline kidney impairment is present in 20% to 35% of patients, and acute kidney injury occurs in 3.3% of patients treated with a contrast agent, especially in those with a high risk owing to baseline kidney impairment.^19^ Moreover, the risk of contrast-induced encephalopathy (CIE), which occurs at a frequency of approximately 1.7% among patients receiving contrast agents, is higher among patients with kidney dysfunction and a history of stroke.^20^ The reported incidence of CIE is 6.8% in patients with chronic kidney disease and 37.5% in those with kidney failure with replacement therapy. To prevent CIE, patients at risk must be identified, the use of high-dose contrast media must be minimized, and alternative imaging modalities should be used when possible.^21^

In the future, the generalizability of the model should be verified in larger, prospective studies, and more clinical observational studies should be conducted, incorporating data such as age, sex, syndrome, background diseases, etc.

This study has some limitations. First, the sample was relatively small, and caution must be exercised in the interpretation and clinical application of the study results. This study was based on data from non-consecutive cases from several regional CSCs in Japan, and bias in the patient population cannot be ruled out. Second, the observer experiment in this study did not faithfully reflect the clinical diagnostic workflow. As it was performed with NCCT readings without clinical information (NIHSS score, symptom side, age, sex, presence of atrial fibrillation, and so on), it may not fully reflect the effect of the model on readings in the real world. Third, our primary focus was on LVO in the anterior circulation and did not include cases of LVO in the posterior circulation.

In conclusion, we developed a fully automated CNN model, tailored to the detection of LVO in patients with anterior-circulation AIS. The CNN can rapidly and reliably detect the HAS on NCCT images and improve neurosurgeons’ diagnostic performance regardless of stroke-care experience.

## Non-standard Abbreviations and Acronyms

AIS: acute ischemic stroke
ASPECTS: Alberta Stroke Program Early Computed Tomography Score
AUC: area under the curve
CIE: contrast-induced encephalopathy
CNN: convolutional neural network
CSC: comprehensive stroke center
CT: computed tomography
CTA: computed tomography angiography
FOM: figure of merit
FROC: free-response receiver operating characteristic
HAS: hyperdense-artery sign
LVO: large-vessel occlusion
MRA: magnetic resonance angiography
MRI: magnetic resonance imaging
NCCT: non-contrast-enhanced computed tomography
NIHSS: National Institutes of Health Stroke Scale
RBC: red blood cell
ROC: receiver operating characteristic

## Author contributions

H.T. designed and conceptualized the study, gathered and analyzed the data, and wrote the initial draft of the manuscript. H.N. and Y.Ab. analyzed the data and revised the manuscript for intellectual content. T.F., A.T., and H.I. performed data analysis and created the automated software. A.I. designed and conceptualized the study, played a major role in data acquisition, and accepts full responsibility for data integrity and accuracy of the data analysis. Y.Ar. has reviewed and approved the study design and the manuscript.

## Acknowledgments

The authors thank Dr. T. Kikuchi, Dr. Y. Yamao, and Dr. Y Terada of the University of Kyoto and Dr. E. Ogino and Dr. R. Goda of Uji Tokushukai Hospital for their cooperation in the observer study.

